# Excess Dietary Fructose Does Not Alter Gut Microbiota or Permeability in Humans: A Randomized Controlled Pilot Study

**DOI:** 10.1101/2020.11.23.20235515

**Authors:** Jose O. Aleman, Wendy A. Henderson, Jeanne M. Walker, Andrea Ronning, Drew Jones, Peter J. Walter, Scott G. Daniel, Kyle Bittinger, Roger Vaughan, Robert MacArthur, Kun Chen, Jan L. Breslow, Peter R. Holt

## Abstract

**Background and Aims:** Non-alcoholic fatty liver disease (NAFLD) is an increasing cause of chronic liver disease that accompanies obesity and the metabolic syndrome. Excess fructose consumption can initiate or exacerbate NAFLD due in part to a consequence of impaired hepatic fructose metabolism. Pre-clinical data have emphasized that fructose-induced altered gut microbiome, increased gut permeability, and endotoxemia play an important role in NAFLD, but human studies are sparse. The present study aimed to explore the relevance of these pre-clinical studies to observations in humans.

**Methods:** We performed a classical double-blind metabolic unit study in 10 obese subjects (BMI 30-40 mg/kg/m^2^) providing 75gms. of either fructose or glucose in their individual diets substituted isocalorically for complex carbohydrates in a cross-over study. Excess fructose intake was provided in the fructose arm of the study and totaled a mean of 22.7% of calories.

**Results:** Routine blood, uric acid, liver function and lipid measurements were unaffected by the fructose intervention. The fecal microbiome (including *Akkermansia muciniphilia*), fecal metabolites, gut permeability, indices of endotoxemia, gut damage or inflammation and plasma metabolites were essentially unchanged by either intervention.

**Conclusions:** Although pre-clinical rodent studies have shown that excess fructose causes pronounced changes in the gut microbiome, metabolome, and permeability as well as endotoxemia, this did not occur in obese individuals fed fructose in amounts known to enhance NAFLD. Therapeutic efforts to improve NAFLD through changes in the gut microbiome and gut homeostasis may not be beneficial.

## Introduction

Dietary fructose consumption in the United States has risen markedly in the past four decades and along with increased total sugar consumption has been correlated with the increased prevalence of the metabolic syndrome[1]. During the same period, non-alcoholic fatty liver disease (NAFLD) has increased with over 25% prevalence in the adult US population in the context of the obesity epidemic[2]. NAFLD may lead to non-alcoholic steatohepatitis (NASH), cirrhosis and liver cancer[3]. Fructose feeding induces fatty liver disease in rodents [4] and zebrafish [5] and has been shown to trigger or worsen NAFLD in humans[6]. A fructose bolus in humans increases serum triglycerides and palmitate[7] and nine days of a diet including 25% of total calories as fructose increases hepatic fatty acid synthesis[8] which is reversed by substitution of fructose by starch[9]. In monkeys, fructose supplementation leads to hepatic inflammation and hepatic fibrosis[10]. Much of this work has recently been summarized[11]. The current mechanistic hypothesis for these effects focuses on fructose-induced changes in hepatic lipid metabolism[12]. Fructose undergoes 1^st^ pass metabolism in the liver[13] and is a substrate for *de novo* lipogenesis driving triglyceride accumulation which causes cellular injury[14].

However, there is also pre-clinical data suggesting that at least part of the ill effects of excess fructose occur from changes in the gut. Small intestinal fructose absorption is limited compared to glucose absorption. Many individuals cannot absorb more than about 25-50 grams of fructose given as a bolus[15]. Fructose absorption is limited because of selective absorption mechanisms for small intestinal transport of fructose[16]. Unabsorbed fructose passes into the colon where it is rapidly fermented by gut bacteria into short chain fatty acids, hydrogen, carbon dioxide and methane[17]. Thus, large amounts of dietary fructose not absorbed in the small intestine passing into the colon might rapidly alter the distribution and function of colonic microbiota[18] leading to changes in microbial metabolite production. Microbial changes after feeding diets high in fructose content have been described in rats[19]. One consequence of fructose-induced changes in gut microbiota and metabolites in rodents is increased intestinal permeability. In mice, increased fructose feeding alters intestinal tight junctions[20, 21]. Antibiotics or transfer of feces from chow-fed rats into the colon of fructose-fed rats reverses gut microbial changes with amelioration of the metabolic syndrome[19]. Altered fecal microbiota have been described in human fatty liver disease[22], and probiotics may reduce liver fat[23].

Increased intestinal permeability has been described in NASH[24, 25] accompanied by endotoxemia[26]. Such endotoxemia is known to sensitize hepatic Kupfer cells inducing inflammation in the liver[27]. In mice, fructose-induced gut permeability is ameliorated by antibiotics[28]. However, we have found no studies that determine effects of fructose feeding on the gut microbiome and metabolites, nor on microbial translocation or intestinal permeability in humans.

The present study was designed to test the hypothesis that fructose causes changes in gastrointestinal microbiota that might contribute to fructose-induced liver disease. Positive findings may allow for novel therapeutic approaches aimed at the gut to partially mitigate these deleterious changes in the liver.

## MATERIALS AND METHODS

### Subjects

Subjects were recruited from the surrounding community and from the Rockefeller University volunteer database. Eligible were obese men and obese postmenopausal women (BMI 30-39 mg/kg/m^2^) between the ages of 45 and 70 years, without clinical or electrocardiographic evidence of cardiometabolic disease. Exclusion criteria were fasting blood glucose >126mg/dL, HgA1C > 6.5%, serum triglyceride levels > 200mg/dL, liver function tests > 1.5 times the upper limit of normal, serum uric acid level > 9mg/dL, current statin, insulin or oral hypoglycemic agents, daily laxatives, probiotics, or proton pump inhibitor usage. Also excluded were individuals who have a history of broad-spectrum antibiotics therapy during the preceding 45 days, active hepatitis, HIV infection, chronic constipation or diarrhea, inflammatory bowel disease, gastrointestinal resection, or macronutrient malabsorption. In addition, current tobacco smokers, and those with a history of more than 8 grams alcohol intake daily were excluded.

Twenty subjects were screened; thirteen met the enrollment criteria and were randomized to receive dietary glucose or fructose replacement in the first study arm (**Figure 1**) and were admitted to the Rockefeller University metabolic unit. Women were postmenopausal to avoid variation in fecal microbiota that could occur during the menstrual cycle[29]. One subject withdrew for personal issues, and two for elevated fasting blood glucose after enrollment. Of 10 participants completing the study, six were men and four women, 2 Caucasian, 1 Black Hispanic, and 7 African American, self-reported. All participants read and signed an informed consent document approved by the Institutional Review Board of The Rockefeller University (Protocol PHO-0956).

**Figure 1:**
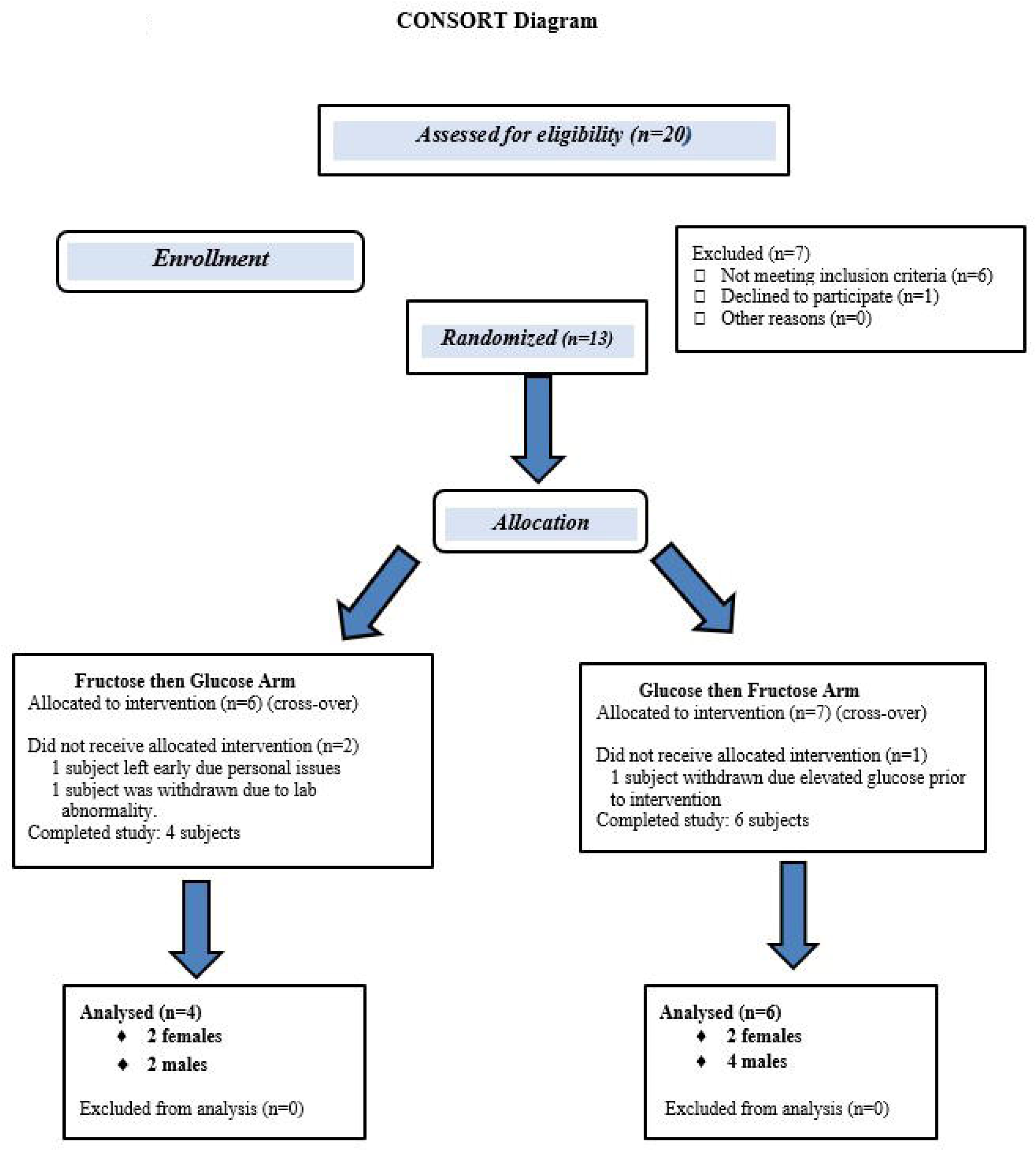
CONSORT Diagram of study participants

### Clinical Study Design and Setting

This was a pilot, double-blind, randomized, cross-over study. Subjects initially were screened in the Outpatient Research Center at the Rockefeller University Hospital. Screening included a complete history and physical examination, fasting blood tests, electrocardiogram, and hip and waist circumference measurements. Subjects met with the bionutritionist for a detailed three-day dietary recall history and completed a computerized diet questionnaire (VioScreen) to determine the participant’s “usual” dietary intake. As fructose is very sweet and can be unpalatable, participants were given taste tests of the fructose solution, mixed in water, lemonade, or tea, to determine which mixture they preferred.

Eligible subjects were randomized by the research pharmacist using SAS 9.4 and the PROC PLAN procedure resulting in 4 subjects receiving fructose replacement first and then glucose replacement, and 6 receiving glucose replacement first followed by the fructose phase. Stratification for gender was balanced within each sequence. To prevent rapid changes in the distribution of fecal microbiota with introduction of a new diet, subjects throughout the study were fed their “usual” diets mimicking diets which they consumed prior to entering the hospital. The daily 75 grams of study sugar was accompanied by removal of 75 grams of complex carbohydrate from the “usual” diets.

Subjects were admitted to the inpatient metabolic unit of The Rockefeller University Hospital for 16-18 days in each study arm (**Figure 2**). They were fed their “usual” diet for the first 2-3 days while undergoing baseline testing including body composition measurement. Fecal samples were collected for microbiome, metabolome, and calprotectin assays followed by a 4-sugar test of gut permeability. Subjects then started the 14-day fructose or glucose diet arm of the study. Subjects, care providers, and investigators except for the study clinician, were blinded to the assigned intervention, and the study drinks were dispensed by the pharmacist in identical receptacles.

**Figure 2:**
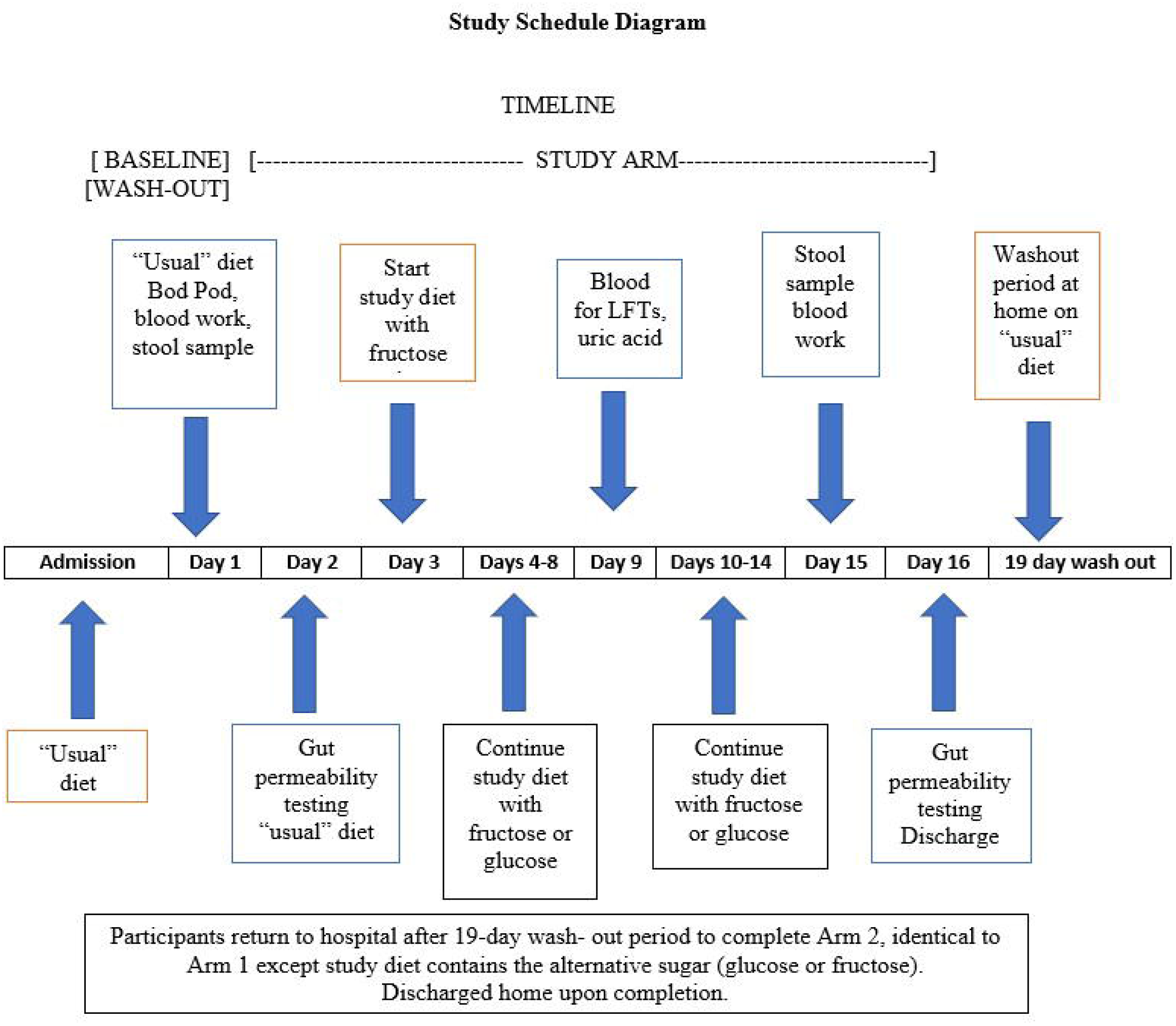
Study schedule diagram.

Body weight was monitored daily and kept stable by adjusting caloric intake. Activity was monitored by a New Lifestyles NL-800 accelerometer (New Lifestyles, Lee’s Summit, MO). On non-testing days, participants were permitted to leave the hospital after breakfast with a packed lunch, but then returned for dinner. The glucose/fructose solution was prepared by the research pharmacy and administered during breakfast and dinner. The bionutrition staff ensured that food provided was acceptable and consumed, and any food that was not eaten was recorded.

Seven days after starting the glucose or fructose arm of the study, liver function, serum triglyceride and uric acid levels were monitored as safety measures. At the end of each study arm, the same tests were administered as at the beginning of the study arm (except for the Bod Pod). The subjects returned home for the 19-day wash out-period, maintained their “usual” diet, received weekly phone contact by the bionutritionist to ensure dietary compliance.

### Anthropometric measurements

Body weight was measured daily with a Scale-Tronix 5002 scale (Welch Allyn, Skaneateles Falls, NY) with precision of +/- 0.1kg. Subjects were weighed in a hospital gown, after an overnight fast and post-voiding. Height was measured to the nearest 1 cm at baseline with a Seca-216 stadiometer (Hamburg, Germany) in 1 mm increments. Body mass index (BMI) was calculated as weight/height squared (kg/m^2^), using the NIH Standard Metric BMI calculator.

### Daily blood pressure monitoring

After sitting for five minutes, manual blood pressure readings (Welch Allyn aneroid monitor, Skaneateles Falls, NY) were taken each morning.

### Diet formulation process and analysis of dietary fructose content are provided in Supplemental Material and Methods

#### Body composition

Bod Pod (Cosmed, Rome, Italy) for body composition assessment, is an air displacement plethysmograph which uses whole body densitometry to determine body composition (fat and fat-free mass).

#### Blood collection and measurements

Fasting blood samples were analyzed in the Clinical Pathology Laboratory of the Memorial Sloan-Kettering Cancer Center for complete blood count, electrolytes, glucose, creatinine, BUN, AST, ALT, alkaline phosphatase, C-reactive protein, sedimentation rate, and uric acid. Research serum and plasma samples were drawn pre and post intervention, aliquoted and stored at −80°C for subsequent analysis.

#### Bacterial profiling of feces

Immediately after defecating, bean sized fecal specimens were collected in triplicate from the middle of the stool, placed in 10 ml Falcon tubes and rapidly frozen at −80°C until analysis. Fecal microbiome profiling was performed at MR DNA (Shallowater, TX) as described in the Supplemental Materials and Methods.

#### Fecal and plasma metabolomics

Fecal and plasma metabolomics were performed at the New York University Metabolomics Core Resource Laboratory as previously described[31]. Samples were subjected to LCMS analysis to detect and quantify known peaks (Supplemental Materials and Methods).

### Fecal fructose and glucose measurements are described in Supplemental Material and Methods

#### Surrogate markers of microbial translocation and gut inflammation

Circulating proteins associated with an increased plasma lipopolysaccharide content were assessed by measuring lipopolysaccharide-binding protein, (Cell Sciences, Inc., Canton, MA) and soluble CD14 concentrations (R&D Systems, Inc., Minneapolis, MN) via an ELISA following the manufacturer’s instructions. Serum concentration of intestinal fatty acid binding protein (Hycult Biotech, Inc., Wayne, PA) and fecal calprotectin (Genova Diagnostics, Ashville, NC) also were detected by an ELISA following the manufacturer’s instructions and were used as markers of gut damage and inflammation respectively. All measurements were performed in duplicate.

#### Gastrointestinal permeability

Gastrointestinal permeability was determined as previously described[30]. The lactulose/ mannitol ratio is used to assess small intestinal permeability and sucralose and sucrose are used to assess colonic and gastric permeability, respectively. In brief, after a baseline urine collection, fasting subjects drank a permeability test solution (100 ml solution containing sucrose [10 g/dl], lactulose [5 g/dL], mannitol [1 g/dL], and sucralose [0.1 g/dL]) and urine was collected for 5 hours. Urine was stored at −80°C until analysis. Analysis of sucralose, sucrose, lactulose, and mannitol in urine was performed in duplicate as described in the Supplemental Materials and Methods.

### Statistical Analysis

#### Sample size analysis

The primary statistical analysis compares the changes in fecal microbiota distribution between two diets supplemented with fructose or glucose. A cross-over experiment submitted each subject to both diets in a randomized order. A sample of *n*=6 would allow 80% power to detect minimum differences as 2-fold changes in the fecal microbiota distribution at 5% significance with the use of Wilcoxon non-parametric tests. This power analysis is based on a variability that is 20% inflated when compared to the average coefficient of variation (∼ 40%) reported by DiLucca et al.[19] when comparing microbiota distribution between two groups of six different rats each. All calculations were performed with G* power software version 3.1.

## RESULTS

Participants who successfully completed the study were 57.6 +/-6.2 years of age. They were obese, weighing an average of 101.8 kg (BMI 35.9 +/-3.3), with a mean body fat composition of 41.1% (**Table 1**). Three subjects in the fructose arm and one subject in the glucose arm reported abdominal discomfort following the sugar drink. Participants’ activity was similar as judged by pedometer step counts (mean for the fructose and glucose arms: 9523 steps and 9124 steps per day, respectively).

**Table 1.** Participant Characteristics

| **Participant Characteristics** |  |  |
| --- | --- | --- |
| **Measurement** | **Mean (SD)** | **Range** |
| *N=10* |  |  |
| Age (years) | 57.6 (6.2) | 50-67 |
| BMI (kg/m<sup>2</sup>) | 35.9 (3.3) | 30.9-39.9 |
| Weight (kg) | 101.8 (14.8) | 80.5-119 |
| Waist (cm) | 114.3 (0) | 98.5-130 |
| Hips (cm) | 119.2 (9.6) | 101-133 |
| Fat Free Mass (%) | 58.6 (6.9) | 45.6-69.4 |
| Fat Mass (%) | 41.4 (6.9) | 30.6-54.4 |

BMI: Body mass index = weight in kilograms/height in meters squared.
Fat mass measured by Bod Pod.

The calculated “usual” dietary intake contained a mean of 3258 kcal with 54% of calories as carbohydrates, 15% as protein, and 33% as fat (**Table 2**). The caloric density and distribution of macronutrients in the designed study diets and the actual consumed diets were very similar to the subjects’ “usual” diets.

**Table 2.**
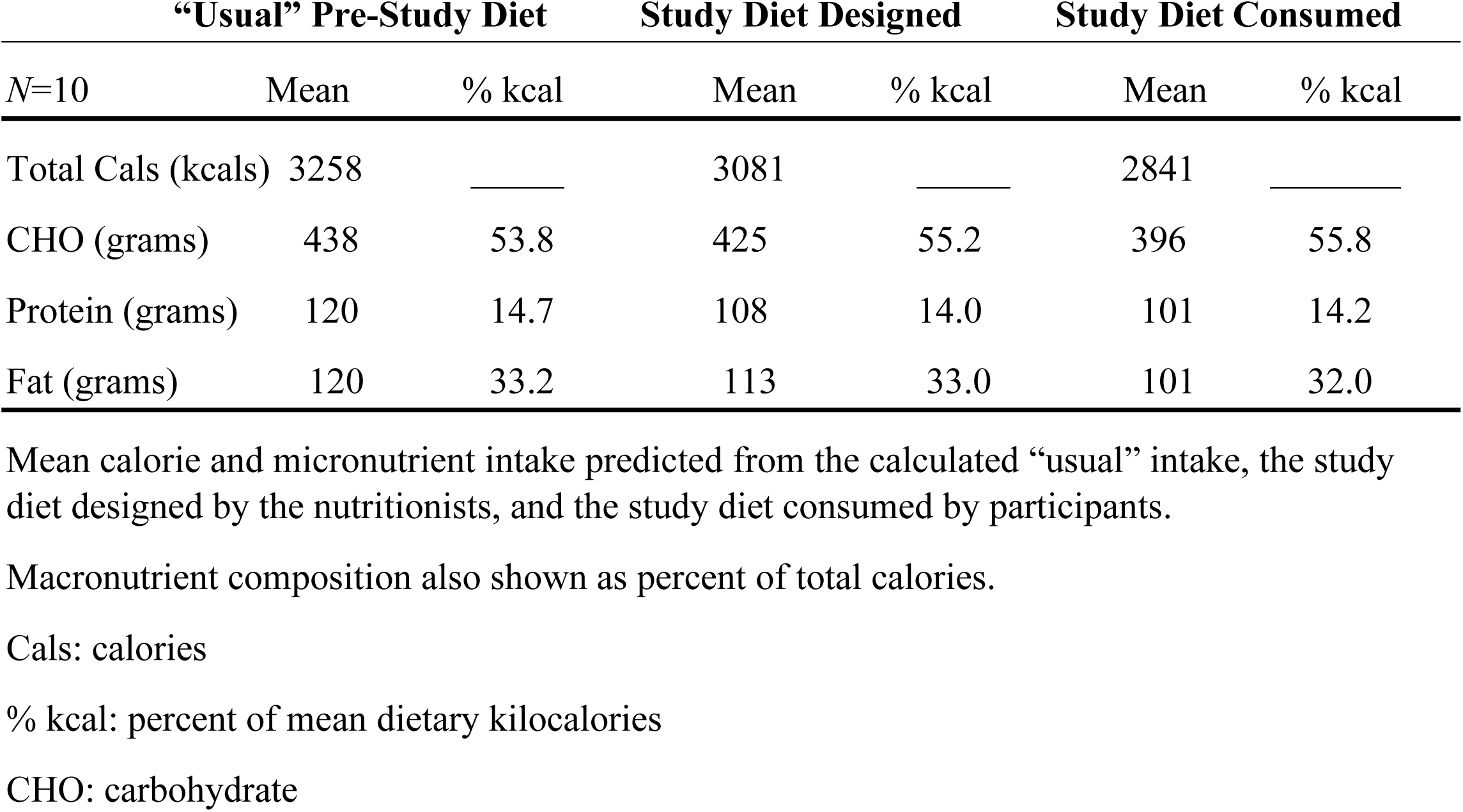
Predicted and Consumed Calorie and Macronutrient Intake

|  | **“Usual” Pre-Study Diet** |  | **Study Diet Designed** |  | **Study Diet Consumed** |  |
| --- | --- | --- | --- | --- | --- | --- |
| *N*=10 | Mean | % kcal | Mean | % kcal | Mean | % kcal |
| Total Cals (kcal) | 3258 | _____ | 3081 | _____ | 2841 | _____ |
| CHO (grams) | 438 | 53.8 | 425 | 55.2 | 396 | 55.8 |
| Protein (grams) | 120 | 14.7 | 108 | 14.0 | 101 | 14.2 |
| Fat (grams) | 120 | 33.2 | 113 | 33.0 | 101 | 32.0 |

Mean calorie and micronutrient intake predicted from the calculated “usual” intake, the study diet designed by the nutritionists, and the study diet consumed by participants.
Macronutrient composition also shown as percent of total calories.
Cals: calories
% kcal: percent of mean dietary kilocalories
CHO: carbohydrate

### Analysis of routine clinical blood indices, microbial translocation, and gut inflammation

No significant differences between the 2 arms of the study were noted in the routine hematology or biochemistry values including lipids, AST, ALT, and uric acid levels. Also, no significant differences in any of the surrogate markers of microbial translocation or gut inflammation were found between the 2 arms. These included serum levels of soluble CD14 and lipopolysaccharide binding proteins as surrogate markers for changes in circulating lipopolysaccharide concentrations (**Table 3**). Fecal calprotectin and serum levels of intestinal fatty acid binding protein as measures of intestinal inflammation and damage respectively showed no changes between the fructose or glucose arms of the study. All results were within the normal range.

**Table 3.**
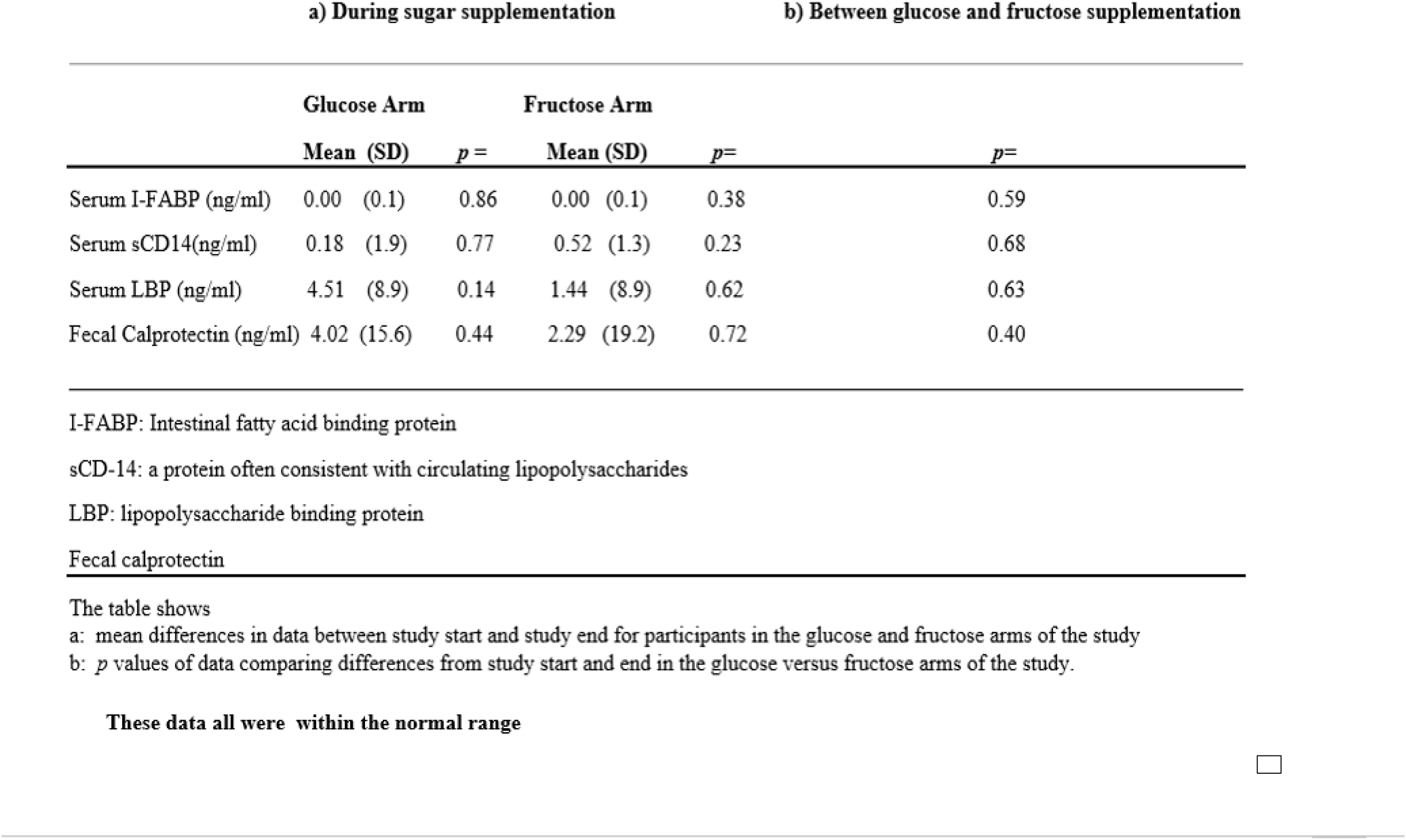
Markers of Gut Translocation and Inflammation (*N*=10)

### Fecal microbiota and metabolome

Fecal samples were analyzed for microbial composition before and after each study arm using 16S rRNA marker gene sequencing to quantify the relative abundance of bacterial taxa, which yielded a median of 63,207 and a minimum of 39,480 reads per sample. Analysis of 16S marker gene data showed no differences between groups before each treatment in alpha diversity, UniFrac distance, or taxonomic abundance in the cross-over study (**Supplemental Figure 1**). The gut microbiota exhibited no differential response to glucose or fructose treatment in alpha diversity (**Figure 3A**), as measured by richness (*P*=0.32), Shannon diversity (*P*=0.43), and Faith’s phylogenetic diversity (*P*=0.47). Beta diversity also was not different between the treatments (**Figure 3B**), based on unweighted UniFrac (*P*=0.99) and weighted UniFrac distances (*P*=1.0). Differences in taxon relative abundance, was sought from all genus-level bacterial taxa with a mean abundance of more than 1% (**Figure 3C**). Based on a mixed effects model of logit-transformed relative abundance, *Bifidobacterium* decreased after treatment in both groups (*P*=0.004) but did not different between treatments (*P*=0.89). *Bifidobacterium* was the only taxon that showed a statistically significant change in abundance after correction for multiple comparisons. In summary, we observed no microbiota associations that differed between treatments.

**Figure 3:**
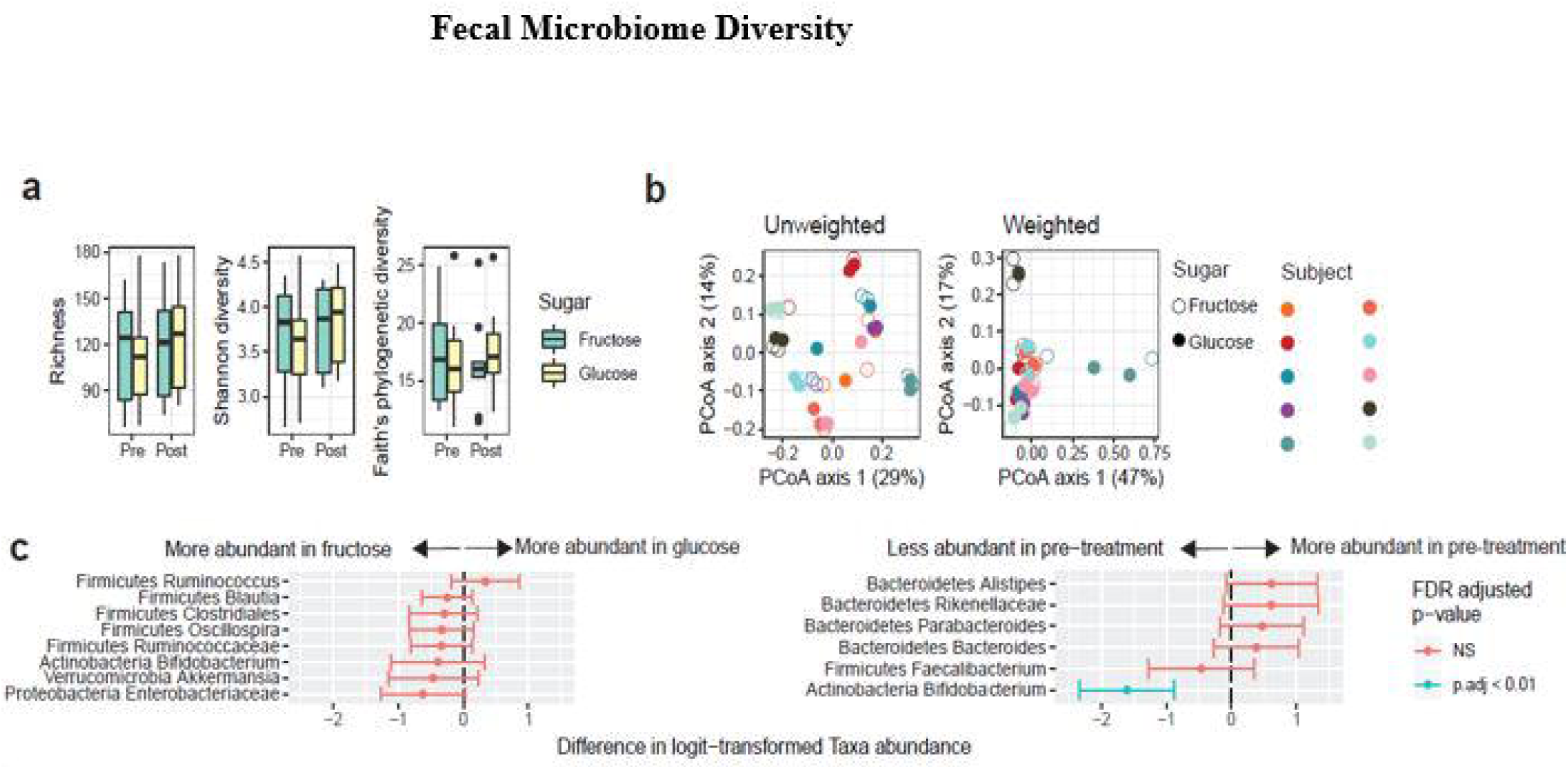
Isocaloric fructose administration preserves fecal microbiome diversity. **a**) alpha diversity (richness calculated at a rarefaction level of 1,000 OTU’s), **b**) beta diversity (unweighted and weighted Unifrac distance), individual subjects shown by different colors on the right and **c**) changes in abundance of specific taxa (outcome variable of logit-transformed abundance based on a mixed-effects model of the predictive variables: type of sugar and pre / post treatment).

Specific analysis of *Akkermansia muciniphila* and *Lactobacillus johnsonii*, which previously had changed with fructose treatment in mice[28], was performed by aligning the unique 16S rRNA amplicon sequence variants in our dataset to the 16S rRNA gene sequence of the type strains for each species to identify their closest representative. No differences in relative abundance for *Akkermansia muciniphilia* and *Lactobacillus johnsonii*, using a mixed effects model were found.

### Fecal metabolome

Overall, few significant metabolite differences were observed in the fructose versus glucose arm following correction for multiple comparisons. To account for inter-personal variation pairwise analysis of data from each subject was performed before and after each treatment arm showing significantly lower levels of fecal glucose and fructose 6-phosphate after fructose supplementation and higher levels of niacinamide and guanine (**Supplemental Figure 2**).

### Fecal fructose

Fecal fructose content showed wide variation between subjects. The study-end fecal specimens contained 36% higher relative peak intensities than the baseline specimens in the fructose arm of the study, whereas fecal fructose fell by 16% in the glucose arm of the study (**Supplemental Figure 3**).

### Plasma metabolome

To gain further insight into the relationship of fecal microbiota, metabolites and changes in intestinal permeability, plasma metabolites were measured at baseline, and study end for each intervention arm. Overall, 115 metabolites were detected in at least one sample, and 66 of these metabolites were detected in all 40 samples. These data were evaluated by principal components analysis, unsupervised hierarchical clustering, and volcano plots and few significant (*p*<0.05) differences were observed using the group-wise statistical comparisons. Similarly, the overall metabolite profile was unable to distinguish the four groups, with all samples appearing equally dispersed. Pairwise analysis using the DESeq2 genomics library[33] to model subject covariate and calculate a corrected *p*-value showed no individual metabolite significantly differed in the glucose arms but methionine increased by ∼7% (*p-*adj=0.035) and ornithine decreased by 1.8-fold (*p*-adj=0.035) in the fructose arm.

### Intestinal permeability studies

To study the effects of fructose compared to glucose feeding on gut permeability, a four-sugar solution was administered before and after the 14-day sugar supplemental diets. No significant differences in urinary excretion of the test sugars used to detect increased permeability in the stomach, small, or large intestine were observed (**Figure 4**).

**Figure 4:**
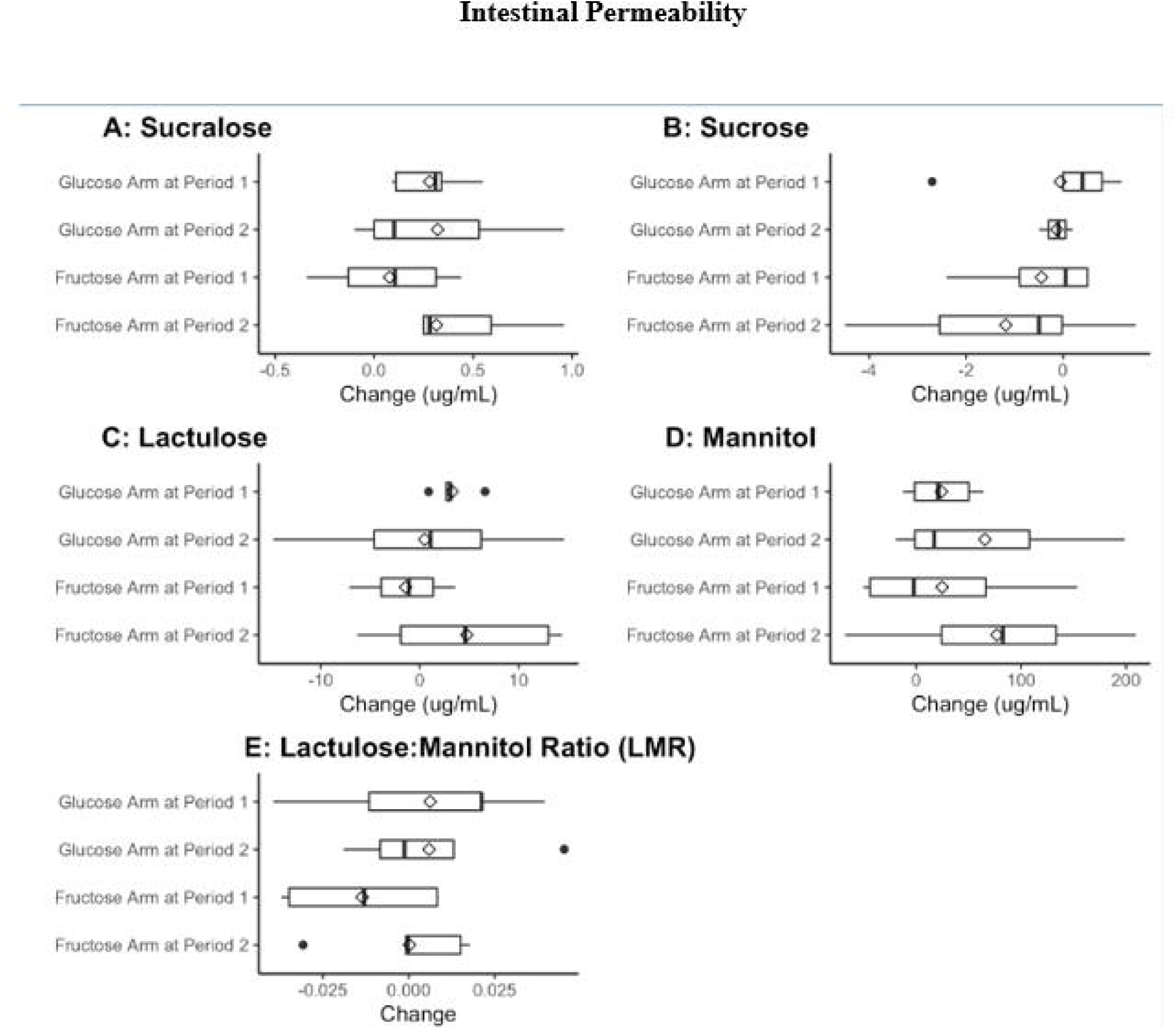
Intestinal permeability is unchanged by fructose administration. Quantification of intestinal permeability by ultra-performance liquid chromatography mass spectrometry. Analysis of changes in concentration of sucrose, sucralose, lactulose, and mannitol in urine. Cross-over design analysis of both diet and timing of the fructose or glucose arm of the study.

We then conducted a crossover design analysis to assess both the diet differences and the timing of the fructose or glucose arm. Again, no significant differences were detected in these test sugars (*p*>0.05). Since residual analysis showed potential outliers, we repeated analysis after removing outliers, but differences in diets or timing remained insignificant. Based on the data from our 10 subjects we would need to recruit 36-182 participants to find a significant difference in urinary sugar excretion between the fructose and glucose arms (Supplemental Materials and Methods).

## DISCUSSION

The present study aimed to determine the effects of excess fructose compared to glucose ingestion on the fecal microbiome, metabolome, intestinal permeability, and markers of endotoxemia in obese individuals in a rigorous cross-over study conducted in an inpatient metabolic unit. Since bacterial fructose metabolism likely impacts colonic homeostasis, we needed to provide enough dietary fructose to ensure that significant amounts of the sugar entered the colon from the small intestine. This goal was achieved as the fecal content of fructose increased by 36% during fructose supplemental feeding but fell by 16% during the glucose supplemental diet. Since fructose is rapidly metabolized by colonic bacteria, the finding of excess fructose in the feces indicated that considerable fructose passed into the colon. The dietary regimen used achieved a fructose intake of as much as a mean of 22.7 % of total calorie intake. The average fructose consumption in the United States population has been calculated as about 10% of calories[34], noting that frequent consumption of sugary drinks can add 9% or more of fructose as percent of calories[35], a total figure that approaches amounts used in our study. We also examined changes in fecal microbiome distribution and function following the fructose-glucose regimens. Switching subjects from their normal diet to a standard metabolic study diet would be expected to rapidly change gut microbiota[18]. To eliminate such effects, we determined and mimicked details of each subject’s “usual” diet as we have previously reported[36], modifying these diets only by reducing complex carbohydrates for the fructose or glucose supplemental drinks to maintain macronutrient stability. These goals were achieved since the planned study diets and the macronutrient content consumed by subjects during the study was very close to their “usual” diets.

Neither the fructose nor glucose diets significantly changed fasting routine laboratory blood tests, liver function tests or lipid and uric acid levels taken 10-12 hours after the last meal. Changes in postprandial lipids are well known to occur after test meals with high fructose content[37], but fasting levels usually are unchanged[38]. Furthermore, no differences in abdominal symptoms between the 2 study periods occurred. Pronounced abdominal symptoms occur following ingestion of a bolus of fructose[39], but the co-presence of dietary glucose increases small intestinal fructose absorption[40] reducing rapid passage of unabsorbed fructose into the colon. Despite this, a considerable amount of fructose must have entered the colon to be metabolized by gut bacteria as fecal fructose increased during fructose supplementation.

Overall, feeding neither the fructose nor glucose supplemented diets for 14 days significantly changed fecal microbial alpha or beta diversity and using untargeted microbial analysis, no significant differences were detected in individual taxa. These data contrast with those described in several studies in the mouse[28, 41, 45].

Furthermore, *Akkermansia muciniphilia* and *Lactobacillus johnsonii* are significantly altered by fructose administration in mice but were not significantly changed by either fructose or glucose feeding in our human subjects. Our data supports others that demonstrates fecal microbiome studies in mice often are not reproduced in human studies[42]. We provided our human subjects a mean of 22.7 percent of total calories as fructose, not significantly less than the 30 percent fed to rodents in several studies [19, 28, 45, 46] or to monkeys [10]. Other investigators gave rodents as much as 50 percent or more fructose in the diet [21, 44).

Although we detected no significant effects on fecal microbiota distribution and individual taxa, these bacteria might have changed their metabolic activity. However, few changes in fecal metabolites were found in our study. In the fructose arm, there were significant reductions in fecal glucose and fructose 6-phosphate levels, presumably representing carbohydrate metabolic re-programming of the existing microbiota. Higher levels of nicotinamide and guanine after fructose were found in the feces. Dietary and bacterial tryptophan are metabolized to niacin by many organisms[43] which also can synthesize purine nucleotides.

In contrast to our human studies, fructose supplemented diets in rodents show changes in gut microbiota[28, 44], and evidence of colonic inflammation[45], increased intestinal permeability[47], and endotoxemia[45], together with changes in gut tight junction enzymes[48]. Our studies showed no evidence of gut damage as judged by fecal calprotectin and circulating intestinal fatty acid binding protein concentrations. Direct measures of intestinal permeability and markers of endotoxemia also were unaltered by either fructose or glucose supplemental feeding.

Pre-clinical studies that show the pronounced changes that excess fructose feeding induces in the gut would imply that they have a role in the initiation or progression of NAFLD. It has been shown that some NAFLD patients may harbor alcohol-producing microbiota in their gut[49, 50] and that probiotics may improve liver function, however, usually associated with some weight loss[51, 52]. Rifaxamin treatment improved liver enzyme and endotoxin concentrations in patients with NAFLD but also lowered body weight[53]. Some children with NAFLD or NASH have a distinctive gut microbiome[54], but the human response to excess fructose is very heterogeneous[55] due to differences in liver metabolism[12]. This might also occur because of varying rates of fructose absorption[16] or differences in fructose microbial metabolism.

We found no detailed studies in the literature on effects of fructose administration on the plasma metabolome. One study reported that fructose feeding increased plasma uridine[56] and another investigator[57] showed fructose fed as high fructose syrup increased plasma lysopholipids and decreased mean acylcarnitine levels suggestive of increased lipid oxidation. Our study showed that fructose compared to glucose administration led to a modest 7% increase in methionine and a 1.8-fold reduction in plasma ornithine concentration which are unlikely to contribute to the ill effects of excess fructose consumption.

A study strength was the cross-over design in a controlled metabolic unit directly comparing equal amounts of fructose or glucose provided in the context of participants’ “usual” diet. The mean of 22.7% of total energy intake as fructose exceed the maximum consumed by humans and was sufficient to increase fructose in the feces implying that considerable fructose reached the colon. A further strength was the simultaneous measures of the fecal microbiome, fecal and plasma metabolites, gut permeability, and markers of endotoxemia, gut damage and inflammation.

A potential weakness was limiting fructose or glucose supplementation to only 14 days, determined in part by practical considerations of this hospital inpatient study. Available data show that the fecal microbiome (and probably fecal metabolites) can change well within 14 days of initiating a change of diet[18, 58]. Direct-acting damaging agents cause gut damage within a few days, but the speed of changing gut permeability is unknown. Although the number of participants was small, the cross-over design enhanced statistical power. As many as 182 subjects would need to be recruited to be able to detect a significant difference between the fructose and glucose study arms of a cross-over study which clearly was not feasible.

In summary, in a controlled metabolic unit cross-over study of equal amounts of fructose or glucose for 14 days, keeping caloric intake stable, excess fructose did not induce significant changes in the fecal microbiome, fecal or plasma metabolites, gut permeability, markers of endotoxemia, gut damage, or inflammation. These data suggest that therapeutic measures that are aimed to change the gut microbiome to improve the treatment of NAFLD may not be beneficial.

## Supporting information

Fructose supplemental material

## Data Availability

All individual participant data collected during the trial, after de-identification, including text, tables, figures, and supplemental material, will be available after publication to anyone who wishes to access the data. Data are available upon reasonable request to:
Dr Peter R. Holt. ORCID identifier 0000-0002-8469-2766 or
Dr. Jose O. Aleman. ORCID identifier 0000-0003-3753-6717.

## Acknowledgements

We thank volunteers for willingly participating in the study as part of the research team, the research nurses for their professional conduct of study and support of our volunteers nutrition staff for support and assurance of dietary compliance

We thank Andrew Dannenberg and David Montrose for providing background data of their work for our application for grant support.

Ariel Myatt for help with preparation of manuscript and figures

## Funding

This research was supported in part by RUCCTS grant # UL1TR001866 from the National Center for Advancing Translational Sciences (NCATS), National Institutes of Health (NIH) Clinical and Translational Science Award (CTSA) Program

The Sackler Center for Biomedicine and Nutrition at the Rockefeller University 1ZIANR000018, Division of Intramural Research, NIH, Department of Health and Human Services.

American Heart Association 17-SFRN33520045

K08-DK117064, NIH, at New York University Langone Health.

## Author Contributions

JOA, JLB, and PRH designed research; JMW conducted research; WAH, PJW,RM provided essential materials; WAH, RV, KC, DJ, SGD, KB analyzed data and performed statistical analysis; PRH, JLB, JMW wrote paper. PRH had primary responsibility for final content; AR designed and prepared study diet. All authors read and approved the final manuscript.

## References

[1] Rodriguez LA, Madsen KA, Cotterman C, Lustig RH. Added sugar and metabolic syndrome in US adolescents: Cross-sectional analysis of the National Health and Nutrition Examination Survey Public Health Nutrition. Public Health Nutr 2016;19:2424–34.

[2] Younossi ZM, Koenig AB, Abdelatif D, Yousef F, Henry L, Wymer M. Global epidemiology of nonalcoholic fatty liver disease-Meta-analytic assessment of prevalence, incidence, and outcomes. Hep 2016;64:73–84.

[3] Calzadilla Bertot L, Adams LA. The natural course of non-alcoholic fatty liver disease. Int J Mol Sci 2016;17: E774.

[4] Stanhope KL, Schwarz JM, Havel PJ. Adverse metabolic effects of dietary fructose: results from recent epidemiological, clinical, and mechanistic studies. Curr Opin Lipidol 2013;24:198–206.

[5] Toop CR, Gentili S. Fructose beverage consumption induces a metabolic syndrome phenotype in the rat: a systematic review and meta-analysis. Nutrients 2016;8:577.

[6] Chen B, Zheng YM, Zhang JP. Comparative study of different diets induced NAFLD models of zebrafish.Front Endocrinol (Lausanne) 2018;9:366.

[7] Hudgins LC, Parker TS, Levine DM, Hellerstein MK. A dual sugar challenge test for lipogenic sensitivity to dietary fructose. J Clin Endocrinol Metab 2011;96:861–8.

[8] Schwartz JM, Noworolski SM, Wen MJ. Effect of a high-fructose weight-maintaining diet on lipogeneses and liver fat. J Clin Endocrinol 2015;100:2434.

[9] Schwartz JM, Noworoloski SM, Erkin-Cakmak A, Korn NJ, Wen MJ, Tai VW, Jones GM, Palii SP, Velasco-Alin M, Pan K et al. Effects of dietary fructose restriction on liver fat, de novo lipogenesis and insulin kinetics in children with obesity. Gastroenterology 2017;153:743–752.

[10] Kavanagh K, Wylie AT, Tucker KL, Hamp TJ, Gharaibeh RZ, Fodor AA, Cullin JM. Dietary fructose induces endotoxemia and hepatic injury in calorically controlled primates. Am J Clin Nutr 2013;98:349–57.

[11] Alwahsh SM, Gebhardt R. Dietary fructose as a risk factor for non-alcoholic fatty liver disease (NAFLD). Arch Toxicol 2017;91:1545-63. [Published on-line 19 December 2016].

[12] Esler WP, Bence KK. Metabolic targets in nonalcoholic fatty liver disease. Cellular Mol Gastroenterol Hepatology 2019;8:247–267.

[13] Havel PJ. Dietary fructose: Implication for dysregulation of energy homeostasis and lipid/carbohydrate metabolism. Nutr Rev 2005;63:133–57. [First published 28 June 2008].

[14] Stanhope KL, Schwarz JM, Keen NL, Griffen SC, Bremer AA, Graham JL, Hatcher B, Cox CL, Dyachenko A, Zhang W et al. Consuming fructose-sweetened, not glucose-sweetened, beverages increase visceral adiposity and lipids and decreases insulin sensitivity in overweight/obese human. J Clin Inves 2009;119:1322–34.

[15] Rao SS, Attaluri A, Anderson L, Stumbo P. Ability of the normal human small intestine to absorb fructose: Evaluation by breath testing. Clin Gastroenterol Hepatol 2007;5:959–63.

[16] Jones HF, Butler RN, Brooks DA. Intestinal fructose transport and malabsorption in humans. Am J Physiol Gastrointest Liver Physiol 2011;300:G202–206.

[17] Gibson PR, Newnham E, Barrett JS, Shepherd SJ, Muir JG. Review article: Fructose malabsorption and the bigger picture. Aliment Pharmacol Ther 2007;25:349–63.

[18] David LA, Maurice CF, Carmody RN, Gootenberg DB, Button JE, Wolfe BE, Ling AV, Devlin S, Varma Y, et al. et al Diet rapidly and reproducibly alters the human gut microbiome. Nature 2014;505:559–63.

[19] DiLucca B, Crecenzo R, Mazzoli A, Cigliano L, Venditti P, Walser J-C, Widmer A, Baccigalupi L, Ricca E, Iossa S. Rescue of fructose-induced metabolic syndrome by antibiotics or fecal transplantation in a rat model of obesity. PLoS One 2015 Aug 5;10(8):e0134893. doi: 10.1371/journal.pone.0134893.

[20] Spruss A, Kanuri G, Stahl C, Bischoff SC, Bergheim I. Metformin protects against the development of fructose-induced steatosis in mice: Role of the intestinal barrier function. Lab Invest 2012;92:1020–32.

[21] Kawabata K, Kanmura S, Morinaga Y, Tanaka A, Makino T, Fujito T, Arima S, Sasaki F, Nasu Y, Tanoue S et al. et al. A high-fructose diet induces epithelial barrier dysfunction and exacerbates the severity of dextran sulfate sodium-induced colitis. Int J Mol Med 2019;43:1487–96.

[22] Boursier J, Mueller O, Barret M, Machado M, Fizanne L, Araujo-Perez F, Guy CD, Seed PC, Rawls JF, David LA et al. The severity of nonalcoholic fatty liver disease is associated with gut dysbiosis and shift in the metabolic function of the gut microbiota. Hepatology 2016; 63: 764–76.

[23] Ahn SB, Jun DW, Kang BK, Lim JH, Lim S, Chung MJ. Randomized, double-blind placebo-controlled study of a multispecies probiotic mixture in nonalcoholic fatty liver disease. Sci Rep 2019;9:1–9.

[24] Wigg AJ, Roberts-Thompson IC, Dymock RB, McCarthy PJ, Grose RH, Cummins AG. The role of small intestinal bacterial overgrowth, intestinal permeability, endotoxemia and tumor necrosis factor alpha in the pathogenesis of non-alcoholic steatohepatitis. Gut 2001;48:206–11.

[25] Luther J, Garber JJ, Khalili H, Dave M, Bale SS, Jindal R, Motola DL, Luther S, Bohr S, Jeoung SW et al. Hepatic injury in nonalcoholic steatohepatitis contributes to altered intestinal permeability. Cell Mol Gastroenterol Hepatol 2015;1:222–32.

[26] Harte AL, DaSilva NF, Creely SJ, McGee KC, Billyard T, Youssef-Elabd EM, Tripathi G, Ashour E, Adballa MS, Sharada HM et al. Elevated endotoxin levels in non-alcoholic fatty liver disease. J Inflamm 2010;7:15.

[27] Chiu C-C, Ching Y-H, Li Y-P, Liu J-Y, Huang Y-W, Yang S-S, Huang W-C, Chuang H-L. Nonalcoholic fatty liver disease is exacerbated in high-fat diet-diet gnotobiotic mice by colonization with the gut microbiota from patient with nonalcoholic steatohepatitis. Nutrients 2017;9:1220.

[28] Volynets V, Louis S, Pretz D, Lang L, Ostaff MJ, Wehkamp J, Bischoff SC. Intestinal barrier function and the gut microbiome are differentially affected in mice fed a western-style diet or drinking water supplemented with fructose. J Nutr 2017;147:770–80.

[29] Koren O, Goodrich JK, Cullender TC, Spor A, Laitinen K, Backhed HK, Gonzalez A, Werner JJ, Angenent LT, Knight R et al. Host remodeling of the gut microbiome and metabolic changes during pregnancy. Cell 2012;150:470–80.

[30] Del Valle-Pinero AY, Van Deventer HE, Fourie NH, Martino AC, Patel NS, Remaley AT, Henderson WA. Gastrointestinal permeability in patients with irritable bowel syndrome assessed using a four-probe permeability solution. Clin Chim Acta 2013;418:97–101.

[31] Pacold ME, Brimacombe KR, Chan SH, Rohde JM, Lewis CA, Swier LJYM, Possemato R, Chen WW, Sullivan LB, Fiske BP et al. A PHGDH inhibitor reveals of serine synthesis and one-carbon unit fate. Nat Chem Biol 2016;12:452–58.

[32] Wishart DS, Feunang YD, Marcu A, Guo AC, Liang K, Vazquez-Fresno R, Sajed T, Johnson D, Karu N et al. HMDB 4.0: the human metabolome database for 2018. Nucleic Acids Res 2018;46:D606–D619.

[33] Love MI, Huber W, Anders S. Moderated estimation of fold change and dispersion for RNA-seq data with DESeq2. Genome Biol 2014;15:550.

[34] Vos MB, Kimmons JE, Gillespie C, Welsh J, Blanck HM. Dietary fructose consumption among US children and adults: The Third National Health & Nutrition Examination Survey. Medscape J Med 2008;10:160. [Published on-line 9 July 2008].

[35] Nielsen SJ, Popkin BM. Changes in beverage intake between 1977 and 2001. Am J Prev Med 2004;27:205–10.

[36] Bokulich NA, Battaglia T, Aleman JO, Walker JM, Blaser MJ, Holt PR. Celecoxib does not alter intestinal microbiome in a longitudinal diet-controlled study. Clin Microbiol Infect 2016;22:464-65. [published online 21 Jan 2016] dx.doi.org/10.1016/j.cmi.2016.01.013

[37] Stanhope KL, Havel PJ. Endocrine and metabolic effects of consuming beverages sweetened with fructose, glucose, sucrose, or high fructose corn syrup. Am J Clin Nutr 2008;88:17335–75.

[38] Dolan LC, Potter SM, Burdock GA. Evidence-based review on the effect of normal dietary consumption of fructose on development of hyperlipidemia and obesity in healthy, normal weight individuals. Crit Rev Food Sci Nutr 2010;50:53–84.

[39] Ravich WJ, Bayless TM, Thomas M. Fructose: incomplete intestinal absorption in humans. Gastroenterology 1983;84:26–29.

[40] Gray GM, Ingelfinger FJ. Intestinal absorption of sucrose in man: interrelation of hydrolysis and monosaccharide product absorption. J Clin Invest 1966;45:388–98.

[41] Ferraris RP, Choe J-Y, Patel CR. Intestinal absorption of fructose. Annu Rev Nutr 2018; 38:41–67.

[42] Wan Y, Wang F, Yuan J, Li J, Jiang D, Zhang J, Li H, Wang R, Tang J, Huang T et al. Effects of dietary fat on gut microbiota and faecal metabolites, and their relationship with cardiometabolic risk factors: a 65 month randomised controlled-feeding trial. Gut 2019;68:1417–29.

[43] Yoshii K, Hosomi K, Sawane K, Kunisawa J. Metabolism of dietary and microbial Vit B family in regulation of host immunity. Front Nutr 2019;17:1–12.

[44] Do MH, Lee E, Oh M-J, Kim Y, Park HY. High glucose or fructose diets cause changes of the gut microbiota and metabolic disorders in mice without body weight change. Nutrients 2018;10:1–14.

[45] Cho YE, Kim D-K, Seo W, Gao B, Yoo S-H, Song B-J. Fructose promotes leaky gut, endotoxemia, and liver fibrosis through ethanol-inducible cytochrome P450-2E1-mediated oxidative and nitrative stress. Hepatology 2019. [published online 8 Apr 2019] doi: 10.1002/hep.30652

[46] Li JM, Yu R, Zhang L-P, Wen S-Y, Wang SJ, Zhang X-Y, Xu Q, Kong L-D. Dietary fructose-induced gut dysbiosis promote mouse hippocampal neuroinflammation: a benefit of short chain fatty acids. Microbiome 2019;7:98.

[47] Rahman K, Desai C, Iyer SS, Thorn NE, Kumar P, Liu Y, Smith T, Neish AS, Li H, Tan S et al. Loss of functional adhesion molecule A promotes severe steatohepatitis in mice on a diet high in saturated fat, fructose, and cholesterol. Gastroenterology 2016;151:733–746e12.

[48] Sellmann C, Priebs J, Landmann M, Degen C, Engstler AJ, Jin CJ, Garttner S, Spruss A, Huber O, Bergheim I. Diets rich in fructose, fat or fructose and fat alter intestinal barrier function and lead to the development of nonalcoholic fatty liver disease over time. J Nutr Biochem 2015;26:1183–1192.

[49] Zhu L, Baker SS, Gill C, Liu W, Alkouri R, Baker RD, Gill SR. Characterization of gut microbiomes in nonalcoholic steatohepatitis (NASH) patients: a connection between endogenous alcohol and NASH. Hepatology 2013;57:601–09.

[50] Yuan J, Chen C, Cui J, Lu J, Yan C, Wei X, Zhao X, Li N, Li S, Xue G et al. Fatty liver disease caused by high alcohol producing Klebsiella pneumoniae. Cell Metab 2019;30:675–88.

[51] Sharpton SR, Maraj B, Harding-Theobald E, Vittinghoff E, Terrault NA. Gut microbiota-targeted therapies in nonalcoholic fatty liver disease: a systemic review, meta-analysis, and meta-regression. Am J Clin Nutr 2019;110:139–49.

[52] Koutnikova H, Genser B, Monteiro-Sepulveda M, Faurie J-M, Rizkalla S, Schrezenmeir J, Clement K. Impact of bacterial probiotics on obesity, diabetes, and non-alcoholic fatty liver disease related variables: a systematic review and meta-analysis of randomized controlled trials. BJM Open 2019;9: e017995.

[53] Gangarapura V, Ince AT, Baysal B, Kayar Y, Kilic U, Gok O, Uysal O, Senturk H. Efficacy of rifaximin on circulating endotoxins and cytokines in patients with nonalcoholic fatty liver disease. Eur J Gastroenterol Hepatol 2015;27:840–45.

[54] Schwimmer JB, Johnson JS, Angeles JE, Behling C, Belt PH, Borecki I, Bross C, Durelle J, Goyal NP, Hamilton G et al. Microbiome signatures associated with steatohepatitis and moderate to severe fibrosis in children with nonalcoholic fatty liver disease. Gastroenterology 2019; 157:1109–22.

[55] Hou R, Panda C, Voruganti VS. Heterogeneity in metabolic responses to dietary fructose. Front Genet 2019;31:1–10.

[56] Yamamoto T, Moriwaki Y, Takahashi S, Tsutsumi Z, Yamakita J, Higashino K. Effects of fructose and xylitol on the urinary excretion of adenosine, uridine, and purine bases. Metabolism 1999;48:520–24.

[57] Gonzales-Granera A, Damns-Machado A, Basrai M, Bischoff SC. Changes in plasma acylcarnitine and lysophosphalidylcholine levels following a high-fructose diet: a targeted metabolomics study in healthy women. Nutrients 2018;10:1254.

[58] Wu W-K, Chen C-C, Panyod S, Chen R-A, Wu M-S, Sheen L-Y, Chang S-C. Optimization of fecal sample processing for microbiome study – the journey from bathroom to bench. J Formos Med Assoc2019;118:545–55.

