## Supplementary material for "Excess Dietary Fructose Does Not Alter Gut Microbiota or Permeability in Humans: A Randomized Controlled Pilot Study": Fructose supplemental material

***Diet formulation***

A bionutritionist interviewed subjects at the first screening visit and completed a food frequency questionnaire using the Vioscreen computer software program and then instructed the subject to complete a detailed 3-day recent diet recall. VioScreen (Viocare, Inc., Princeton, NJ) is a web-based interactive and graphical dietary analysis food frequency questionnaire to determine “usual” nutritional intake. It is conducted on a laptop and requires 30-45 minutes to complete. The results of these measures were reviewed with subjects and used to determine the study participant’s average 3-month caloric, macronutrient, and fructose intake. The 3-day diet recall was used to mimic a typical diet that each subject had consumed before the study. Each subject’s diet was unique and was maintained to keep their fecal microbiota as constant as possible while examining the effect of the supplemented fructose or glucose[S1]. The diets were provided initially during the 2-3-day baseline testing period. The nutritionist also constructed a 3-day rotating diet in which 75 grams of complex carbohydrate were reduced for each subject over 3 meals. A drink containing 37.5 grams of pure fructose or glucose was consumed during breakfast and during dinner providing a total of 75 grams of fructose or glucose.

***Analysis of dietary fructose content***

The Nutrition Data Systems for Research (NDS-R) nutrient analysis program, managed by the Nutrition Coordinating Center of the University of Minnesota was used to analyze the fructose content of the 3-day rotating test diet that was provided to each research participant. Serving size and additional ingredients or foods used in preparation were included. The selected nutrients analyzed determined total fructose content of each day’s menu.

***Bacterial profiling of feces***

Briefly, DNA were extracted from fecal samples and the 16S rRNA gene V4 variable region PCR primers 515/806 were used in a single-step 30 cycle PCR using the HotStarTaq Plus Master Mix Kit (Qiagen) under the following conditions: 94 °C for 3 minutes, followed by 28 cycles (5 cycles used on PCR products) of 94 °C for 30 seconds, 53 °C for 40 seconds and 72 °C for 1 minute, followed by a final elongation step at 72 °C for 5 minutes. Sequencing was performed using an Ion Torrent PGM following the manufacturer’s guidelines.

Sequence data was processed using QIIME2[S2]. Read pairs were processed to identify amplicon sequence variants (ASVs) with DADA2[S3]. Taxonomic assignments were generated by comparison to the Greengenes reference database[S4] using the naïve Bayes classifier implemented in scikit-bio[S5]. A phylogenetic tree was inferred from the sequence data using MAFFT[S6].

Alpha diversity was assessed by richness, Shannon index, and Faith’s phylogenetic diversity. The richness for each sample was estimated as the expected number of ASVs at 1000 reads per sample. Similarity between samples was assessed by unweighted and weighted UniFrac distance [PMID 16332807, 17220268]. Principal Coordinates Analysis (PCA), a method of ordination, was used to create a plot of the sample-sample distances.

To study the cross-over effects, first it was established that there was no significant difference between the baselines of Arm 1 and Arm 2 (represented by A and C, respectively). To compare C to A, we first filtered for taxa that are >1% mean abundance across all samples. Then, we constructed a linear model of the remaining taxa’s abundances vs. the timepoint, with the repeated measure of “Subject” being the random effect. We established that there were no significant taxon abundance changes for timepoint C vs A, and therefore constructed a model comparing just the two treatments. We additionally tested for the fructose effect on taxon abundances, with the additional variable of whether the subject began at timepoint A or C, again adding the repeated measure of “Subject” as the random effect. Similar analyses were carried out for the effects of glucose.

***Measurement of fecal and plasma metabolites***

Metabolite extraction was carried out on each sample after scaling the extraction to a measured aliquot for each sample (~20 mg/mL), using 80% methanol containing internal standards. All samples were reconstituted in water for the analysis. The LC column was a Millipore^TM^ ZIC-pHILIC (2.1 x150 mm, 5 μm) coupled to a Dionex Ultimate 3000TM system and the column oven temperature was set to 25^o^C for the gradient elution. A flow rate of 100 μL/min was used with the following buffers: A) 10 mM ammonium carbonate in water, pH 9.0, and B) neat acetonitrile. The gradient profile was as follows: 80-20%B (0-30 min), 20-80%B (30-31 min), 80-80%B (31-42 min). Injection volume was set to 1 μL for all analyses (42 min total run time per injection). MS analyses were carried out by coupling the LC system to a Thermo Q Exactive HFTM mass spectrometer operating in heated electrospray ionization mode (HESI). Method duration was 30 min with a polarity switching data-dependent Top 5 method for both positive and negative modes. Spray voltage for both positive and negative modes was 3.5kV and capillary temperature was set to 320^o^C with a sheath gas rate of 35, aux gas of 10, and max spray current of 100 μA. The full MS scan for both polarities utilized 120,000 resolution with an AGC target of 3e6 and a maximum IT of 100 ms, and the scan range was from 67-1000 m/z. Tandem MS spectra for both positive and negative mode used a resolution of 15,000, AGC target of 1e5, maximum IT of 50 ms, isolation window of 0.4 m/z, isolation offset of 0.1 m/z, fixed first mass of 50 m/z, and 3-way multiplexed normalized collision energies (nCE) of 10, 35, 80. The minimum AGC target was 1e4 with an intensity threshold of 2e5. All data were acquired in profile mode. Further, the order of acquisition was randomized to mitigate sequence effects and other artifacts. Blank controls (processed side-by-side with samples) and instrument standards were analyzed at regular intervals throughout the acquisition.

***Relative quantification of metabolites***

The resulting Thermo^TM^ RAW files were converted to mzXML format using ReAdW.exe version 4.3.1 to enable peak detection and quantification. The centroided data were searched using an in-house python script Mighty skeleton version 0.0.2 and peak heights were extracted from the mzXML files based on a previously established library of metabolite retention times and accurate masses adapted from the Whitehead Institute[S7], and verified with authentic standards and/or high resolution MS/MS spectral manually curated against the NIST14MS/MS[S8] and METLIN[S9] tandem mass spectral libraries. Metabolite peaks were extracted based on the theoretical m/z of the expected ion type e.g., [M+H]+, with a ±5 part-per-million (ppm) tolerance, and a ± 7.5 second peak apex retention time tolerance within an initial retention time search window of ± 0.5 min across the study samples. The resulting data matrix of metabolite intensities for all samples and blank controls was processed with an in-house statistical pipeline Metabolyze version 1.0 and final peak detection was calculated based on a signal to noise ratio (S/N) of 3X compared to blank controls, with a floor of 10,000 (arbitrary units). For samples where the peak intensity was lower than the blank threshold, metabolites were annotated as not detected, and the threshold value was imputed for any statistical comparisons to enable an estimate of the fold change as applicable. The resulting blank corrected data matrix was then used for all group-wise comparisons.

***Measurements of fecal fructose and glucose***

As formula isomers, glucose and fructose can be challenging to separate chromatographically, since potentially confounding isomers (hexoses) may be found in a complex material such as fecal samples. Therefore, authentic standards for fructose and glucose (U-13C labeled) were analyzed alone and spiked into the fecal metabolite extracts from several samples to test for matrix interference in retention time and resolution. The baseline fructose peak was resolved from the glucose peak allowing discrimination of both metabolites.

Peak intensities for fructose and glucose are reported based on a fixed retention time window, and accurate mass [M-H]^-^ ion at 179.0561 *m*/*z*. (**Supplemental Figure 3**). A potential confounder is the presence of other monosaccharide sugars which might interfere with the detection method. The current Human Metabolome Database (v4.0)[32] describes 26 hexose isomers which could be present in human specimens, but quantities were likely very small compared to the fructose and glucose peaks based on spike-in studies.

***Plasma and fecal metabolomics statistics***

T-tests were performed with the Python SciPy (1.1.0) library[S10] to test for differences and generate statistics for downstream analyses. Any metabolite with *p*-value < 0.05 was considered significantly regulated (up or down). Heatmaps were generated with hierarchical clustering performed on the imputed matrix values utilizing the R library heatmap (1.0.12). Volcano plots were generated utilizing the R library, Manhattanly (0.2.0). To adjust for significant covariate effects (as applicable) in the experimental design the R package, DESeq2 (1.24.0) was used to test for significant differences. Data processing for this correction required the blank corrected matrix to be imputed with zeroes for non-detected values instead of the blank threshold to avoid false positives. This corrected matrix was then analyzed utilizing DESeq2 to calculate the adjusted *p*-value in the covariate model.

***Analysis of urinary sucralose, sucrose, lactulose, and mannitol for permeability studies***

Quantification was achieved by ultra-performance liquid chromatography mass spectrometry utilizing a Thermo Scientific Vanquish UPLC with a Thermo Scientific Q-Exactive mass spectrometer with HESI-II electrospray source at 2500V in negative ion mode at 70,000 resolution.	The internal standard solution contained sucralose, sucrose, ^13^C-mannitol, and ^13^C-lactulose. Calibration standard stocks contained a mixture of sucralose (0.01-15 ug/mL), sucrose (0.01-100 ug/mL), lactulose (0.25-500 ug/mL) and mannitol (0.5-500 ug/mL). Calibration standard stocks or urine (50 uL) was mixed with 950 uL internal solution, vortexed and then centrifuged at 4 ℃, 1400 rpm for 15 minutes. The supernatant was transferred to LC-MS vial and maintained at 4 ℃ for analysis. Injection volume was 1 uL.

Separations were performed on a Waters Cortecs HILIC 2.7 µm, 2.1x100mm column (Waters Corp., Milford, MA) maintained at 35 ⁰C. The separation used solvent A (70% H_2_0, 30% ACN, 0.1% NH_3_) and solvent B (20% H_2_0, 80% ACN, 0.1% NH_3_) at a flow rate of 250 µL/min. The gradient was initially at 100% solvent B, decreased to 75% B at 3 min, decreased to 40% B at 3.5 min and maintained for 1 min, then returned to 100% B at 5.0 min until 7.5 min.

Quantitation was based on the M-H *m/z* and retention time. Analyte and the isotopologue internal standard M-H m/z are sucralose (395.0079, 401.0449), sucrose (341.1099, 327.1466) lactulose (341.1099, 353.11492), and mannitol (181.0714, 187.0919, respectively. The calibration curve had a R^2^>0.999 with 1/x weighting, meets FDA LC-MS guidelines for linearity and quantitation.

***Calculation of participant numbers that would be needed to detect significant differences in urinary sugar excretion between the 2 arms of the study***

We estimated the number of participants needed to find a significant difference between the fructose and glucose arms of the study, using the data derived from our study of 10 participants. The desired power was set as 90% and the significance at 0.05 with some outliers excluded to permit reasonable estimates of the expected effects and standard deviations.

For sucralose, 36 participants would be needed in the 2 treatments arms of a cross-over study to detect a treatment difference of 0.16 ug/ml sucralose at a 2 sided 0.05 significance level with 90% power.

For sucrose, 68 participants would be needed in the 2 treatment arms of a cross-over study to detect a treatment difference of 0.21 ug/ml sucrose at a 2 sided 0.05 significance level with 90% power.

For lactulose, 182 participants would be needed in the 2 treatment arms of a cross-over study to detect a treatment difference of 1.50 ug/ml in lactulose at a 2 sided 0.05 significance level with 90% power.

For mannitol, 176 participants would be needed in the 2 treatment arms of a cross-over study to detect a treatment difference of 15.3 ug/ml in mannitol at a 2 sided 0.05 significant level with 90% power.

**Supplemental Figure 1**

**Fecal Microbiome Diversity**

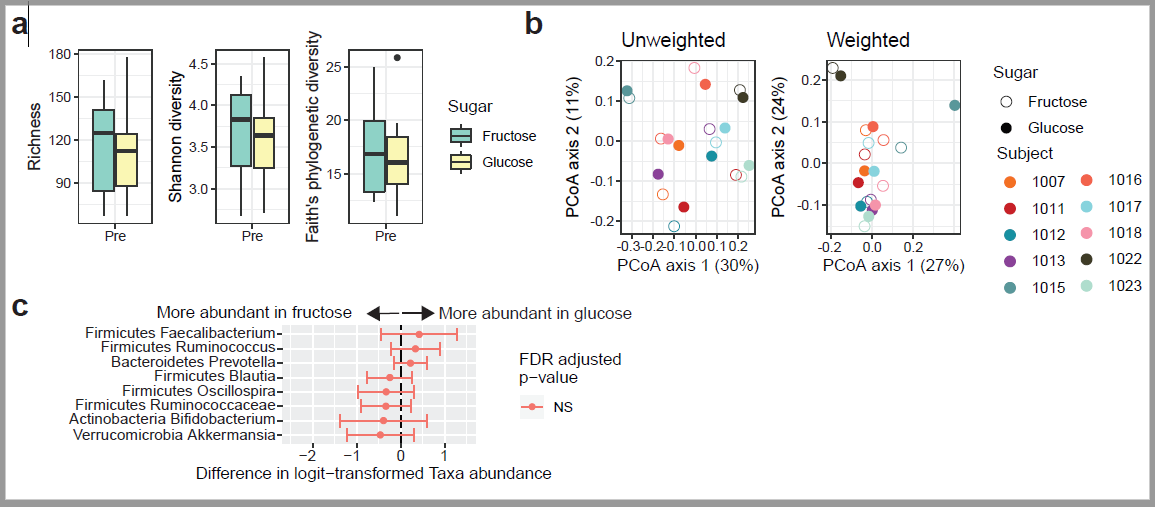

**Supplemental Figure 2	Pairwise Analysis of	Fecal Metabolites**

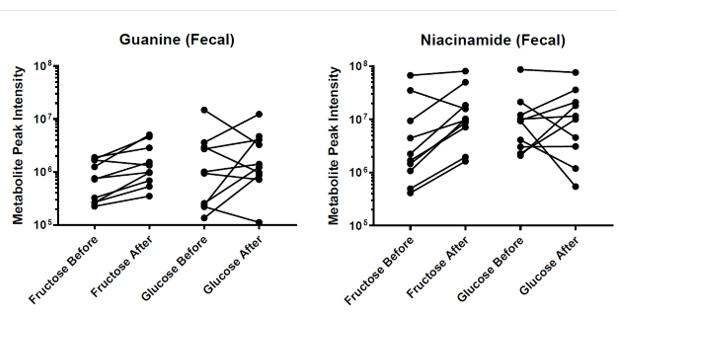

**Supplemental Figure 3**

**Fructose and Glucose Resolution in Fecal Metabolite Extracts**

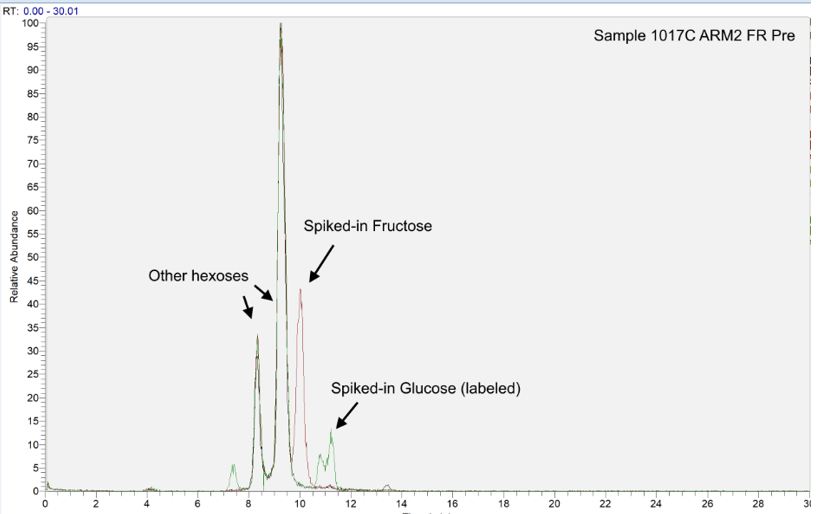

NL: 3.97E6 Base peak m/z= 179.0543-179.0579 F:FTMS-p ESI Full ms (67.0000-10000.0000)MS S03962_H20

NL: 3.95E6 Base peak m/z= 179.0543-179.0579 F:FTMS-p ESI Full ms (67.0000-1000.0000)MS s03962_fructose

NL: 3.80E6 Base peak m/z=179.0543-179+185.0743 – 185.0781 F:FMTS-p ESI Full ms S03962_glucose

**Supplemental Figure Legends**

**Supplemental Figure 1:** Fecal microbiome diversity is unchanged at baseline. For baseline samples only: **a)** alpha diversity (richness calculated at a rarefaction level of 1,000 OTU's), **b)** beta diversity (unweighted and weighted Unifrac distance), individual subjects shown by different colors on the right and **c)** changes in abundance of specific taxa (outcome variable of logit-transformed abundance based on a mixed-effects model of the predictive variable: type of sugar).

**Supplemental Figure 2:** Pairwise analysis of differentially regulated fecal metabolites. Individual values for **Guanine** (left) and **Niacinamide** (right) are plotted, combining the fructose or glucose study arms. Each metabolite value represents detected peak height, shown on a logarithmic scale due to the high inter-personal variation in metabolite profiles. Lines connect metabolite measurements from individual human subjects.

**Supplemental Figure 3**: Authentication of fructose and glucose resolution in fecal metabolite extracts. Three extracted ion chromatograms (XICs) are shown as overlays (fixed intensity) including 1) a representative study fecal sample alone (black), 2) a combined extracted ion chromatogram for Fructose and U-13C-Glucose i.e., Fructose spiked into the sample (red), 3) a combined extracted ion chromatogram for Fructose and U-13C-Glucose i.e., glucose spiked into sample (green).

**Supplemental References**

[S1]	Bokulich NA et al. Celecoxib does not alter intestinal microbiome in a longitudinal diet-	controlled study. *ClinMicrobiolInfect*2016;**22**:464-65.

[S2**]**	Boleyn E et al. Reproducible interactive scalable and extensible microbiome data science	using QIIME2. *NatBiotechnol*2019;**37**:852-57.

[S3]	Callahan BJ et al. DADA2: high resolution sample inference from illumina amplicon data. *NatMethods*2016;**13**:581-83.

[S4]	McDonald D et al. An improved Greengenes taxonomy with explicit ranks for ecological	and evolutionary analyses of bacteria and archaea. *ISMEJl*2012;6:610-18.

[S5]	Bokulich NA et al. Optimizing taxonomic classification of marker-gene amplicon sequences with QIMME 2’s q2-feature-classifier plugin. *Microbiome*2018;**6:**1-17.

[S6]	Kazutaka K, Standly DM. MAFFT multiple sequence alignment software version 7: improvements in performance and usability. *MolBiolEvol*2013;**30**:772-80.

[S7]	Chen WW et al. Absolute quantification of matrix metabolites reveals the dynamics of	mitochondrial metabolism. *Cell*2016;**166**:1324-1337e11.

[S8]	Simon-Manso Y et al. Metabolite profiling of a NIST standard reference material for	human plasma (SRM1950): GC-MS, LC-MS, NMR, and clinical laboratory analysis,	libraries, and web-based resources. *AnalChem*2013;**85**:11725-31.

[S9]	Smith CA et al. METLIN: a metabolic mass spectral database. *TherDrugMonit*2005;**27:**	747-51.

 [S10]	Jones E et al. SciPy: Open Source Scientific Tools for Python, 2001.	 <http://www.scipy.org/> [Online; accessed 2019-08-19].
